# Overweight and its Associated Risk Factors among Students in Tertiary Institutions of Learning in Mongu District of Western Province, Zambia; A cross-sectional Survey

**DOI:** 10.1101/2023.10.12.23296956

**Authors:** Larry L. Mooka, Priscilla Funduluka, Mukumbuta Nawa

## Abstract

**Introduction:** Overweight and obesity are key public health issues in many high-income countries and have become a major public health concern affecting all ages, including adolescents in low-income and middle-income countries. This study determined the prevalence of overweight and obesity and their associated factors among undergraduate students at colleges and universities in the Mongu district of the Western Province of Zambia.

**Methods:** This was a cross-sectional study. Undergraduate students aged 18 to 26 years were sampled from two colleges and one university in Mongu. Data was collected electronically by trained research assistants using a structured questionnaire using Kobo Collect software. Data was analysed using Stata version 14 software. The analysis included descriptive statistics, using counts, frequencies, percentages, means and medians depending on the distribution of the data. Chi-squared tests of association, bivariate, and multivariate logistic regression were done to test for association between overweight and independent variables. A p-value less than 0.05 was considered significant.

**Results:** A total of 330 students were captured in the survey; over half of the respondents were female students 214 (64.85%), while 116 (35.15%) were male respondents. Body Mass Index (BMI) showed that 21 (6.8%) were underweight, 208 (63.0%) had normal weight, 76 (23.0%) were overweight, and 25 (7.6%) were obese. Factors associated with overweight and obesity included female sex aOR 1.68 (95%CI 1.02 – 2.76), age and alcohol intake.

**Conclusions:** A third of the students were either overweight or obese. Sex, age and alcohol intake were significantly associated with overweight and obesity among students. The prevalence of overweight and obesity is high among young adults, and this calls for early interventions in learning institutions to combat obesity.

## BACKGROUND

Obesity is a key public health issue in many high-income countries and is a steadily rising public health problem in low-income and middle-income countries (Almutairi *et al*., 2021). Obesity and overweight have grown to epidemic proportions, with over 4 million people dying each year due to being overweight or obese in 2017, according to the global burden of disease (WHO, 2022).

Overweight and obesity are defined as abnormal or excessive fat accumulation that may impair health (WHO, 2022). Body Mass Index (BMI) is a simple weight-for-height index commonly used to classify overweight and obesity in adults. It is defined as a person’s weight in kilograms divided by the square of his height in meters (kg/m^2^) (WHO, 2022). For adults, a BMI of under 18.5 is considered underweight, a BMI of 18.5 – 24.9 is considered normal weight, a BMI over 25 is considered overweight, and over 30 is obese (WHO, 2022).

The prevalence of overweight and obesity among adults has doubled since 1980 worldwide, and it has increased in many low and middle-income countries (LMICs). Cardiovascular diseases (CVDs), diabetes, musculoskeletal disorders, and some types of cancers associated with overweight and obesity are the leading causes of mortality worldwide (Tateyama *et al*., 2018). Current projections show that the burden of chronic diseases will become comparable to acute infectious diseases in many LMICs (Tateyama *et al*., 2018; Nawa *et al*., 2020).

Sub-Saharan Africa, including Zambia, has experienced an increase in overweight and obesity due to rapid lifestyle changes associated with recent economic growth (Tateyama *et al*., 2018). Obesity-related diseases currently represent 3.8% of disability-adjusted life years (Tateyama *et al*., 2019). The estimated national prevalence of overweight and obesity in Zambia was 29.2% in 2014, compared with 26.4% in 2010 (Bwalya *et al*., 2017; Nawa, 2020).

The increase in overweight and obesity can pose a huge burden on healthcare systems with potential effects on the social and economic well-being of an individual and community. Poor health conditions resulting from overweight and obesity may likewise act as a barrier to developing a good mind, thus paving the way for poor learning behaviour and outcomes among students (Kafyulilo, 2008;).

The study sought to establish the prevalence and to determine the risk factors associated with overweight and obesity among college students in the Mongu District of Western Province, Zambia. While there are few studies on overweight and obesity in Zambia, previous studies have sampled the general population, whilst this study sampled young adults. It will fill the knowledge gap and inform policymakers for this demographic.

## METHODOLOGY

### Study Setting

Mongu is the capital of the Western Province of Zambia and is geographically located 580 kilometres west of Lusaka, the capital city of Zambia. Whilst Western Province is mostly rural, Mongu is the most urbanised and populous area. The people are involved in trade, civil service and agricultural practices. The study was conducted at three (3) tertiary learning institutions: Mongu College of Education, Lewanika College of Nursing and Midwifery and University of Barotseland.

### STUDY DESIGN

This study was a cross-sectional survey conducted between April 2023 and June 2023.

### Study Respondents

The study targeted young adults pursuing tertiary education; this demographic represents young educated people in the province with at least some tertiary education. Therefore, this study will indicate the prevalence and risk factors in the young educated class.

### Inclusion Criteria

All full-time students enrolled at the three selected colleges and universities in the Mongu district were eligible for the study.

### Exclusion Criteria

Pregnant female students, including bodybuilders or weight lifters, were excluded from the study. Students on any form of medication, acutely ill or with known chronic diseases, were also excluded from the study.

## SAMPLE SIZE AND POWER CONSIDERATION

Since the sample for this study was drawn from a finite population, the sample size was determined using the 30% prevalence based on a study done by (Bwalya *et al*., 2017). A precision of 5% (expressed as 0.05) for a 95% confidence level, with a Z-alpha value of 1.96, was used.

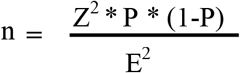

Where:

n = Desired sample

Z = Z-score corresponding to 95% CI

P = Expected prevalence of obesity obtained from previous studies E = Desired precision, which is 5% (expressed as 0.05).

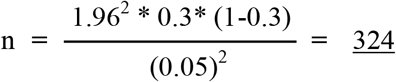

For non-response and refusals, non-response rate was calculated and compensated using the formulae:

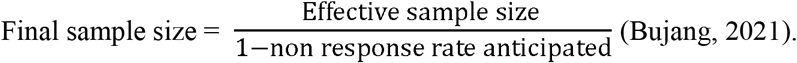

### Sampling Technique

Two-stage stratified random sampling technique was applied in this study because the sample obtained was not homogenous (WHO, 2005). The first-tier sample comprised colleges/universities selected randomly from all the learning institutions in the Mongu district. From the three selected learning institutions, all students who met the eligibility criteria constituted the sampling frame of the study. The second tier was a simple random sampling of the students from the selected learning institutions.

## DATA COLLECTION

Five (5) research assistants were trained to take anthropometric measurements and administer the questionnaire using Kobo Toolbox Collect installed on smartphones (https://ee.kobotoolbox.org/x/hOHLVeP6).

## DATA MANAGEMENT AND ANALYSIS

Data was exported from Kobo Toolbox Collect into Stata version 14.0. Data analysis included computing BMI from the weights and heights (w/h^2^) and categorising BMI into underweight, normal weight, and overweight/obese. The prevalence of the outcome variables (overweight and obesity) was then estimated. Descriptive statistics were presented as frequency, percentages, median and interquartile range were used. Chi-square tests of association were used to analyse associations between categorical variables. Bivariate and multivariate logistic regression analysis was used to assess the associations between the student’s demographics, alcohol intake, food consumption pattern, physical exercise, and eating fruits and vegetables with the outcome. For this study, P≤0.05 was considered statistically significant.

### Ethical Considerations

The study protocol was submitted to the ERES Converge IRB Committee for ethical review and approval (No. 2023-April-007). Permission to conduct the study was sought in writing from each of the three (3) higher learning institutions from which the study was conducted. Written consent was obtained from respondents after explaining to them the purpose of the study and how the results were to be utilised. Data obtained from respondents was kept in a password-protected computer to guarantee respondents’ confidentiality and anonymity. The respondents retained the absolute right and freedom to decline to participate or withdraw from the study at any time without consequence.

### Dissemination Plan

Recommendations have been made to the Ministry of Education regarding implementing overweight/obesity college interventions. Findings will be used to advocate for higher institutions-based curriculum and policy change at the national level. The findings will be published as a Master’s degree dissertation in peer-reviewed publications. In addition, key findings will be shared with the Ministry of Education and participating colleges and universities through report sharing.

## RESULTS

### Socio-Demographic Profile

As shown in Table 1, over half of the respondents were females, 214 (64.85%), while 116 (35.15%) were male respondents. The median weight was 62kg, and the weight IQR=55-70. The majority of the respondents, 106 (32.12%), were within the 21-23-year age group, while the minority of the respondents, 62 (18.79%), were within the 18-20-year age group. A total of 127 (38.48%) students were recruited from Mongu Catholic College of Education (MOCE). In comparison, 126 (38.18%) students were recruited from Lewanika College of Nursing and Midwifery (LCNM) and 77 (23.33%) students from the University of Barotseland (UBL). A widely held percentage of the respondents, 151 (45.76%), were from a non-health-related field, followed by 126 (38.18%) from a nursing field and 53 (16.06%) from a health-related field. A greater proportion of the respondents, 177 (53.64%), were first-year students.

**Table 1:**
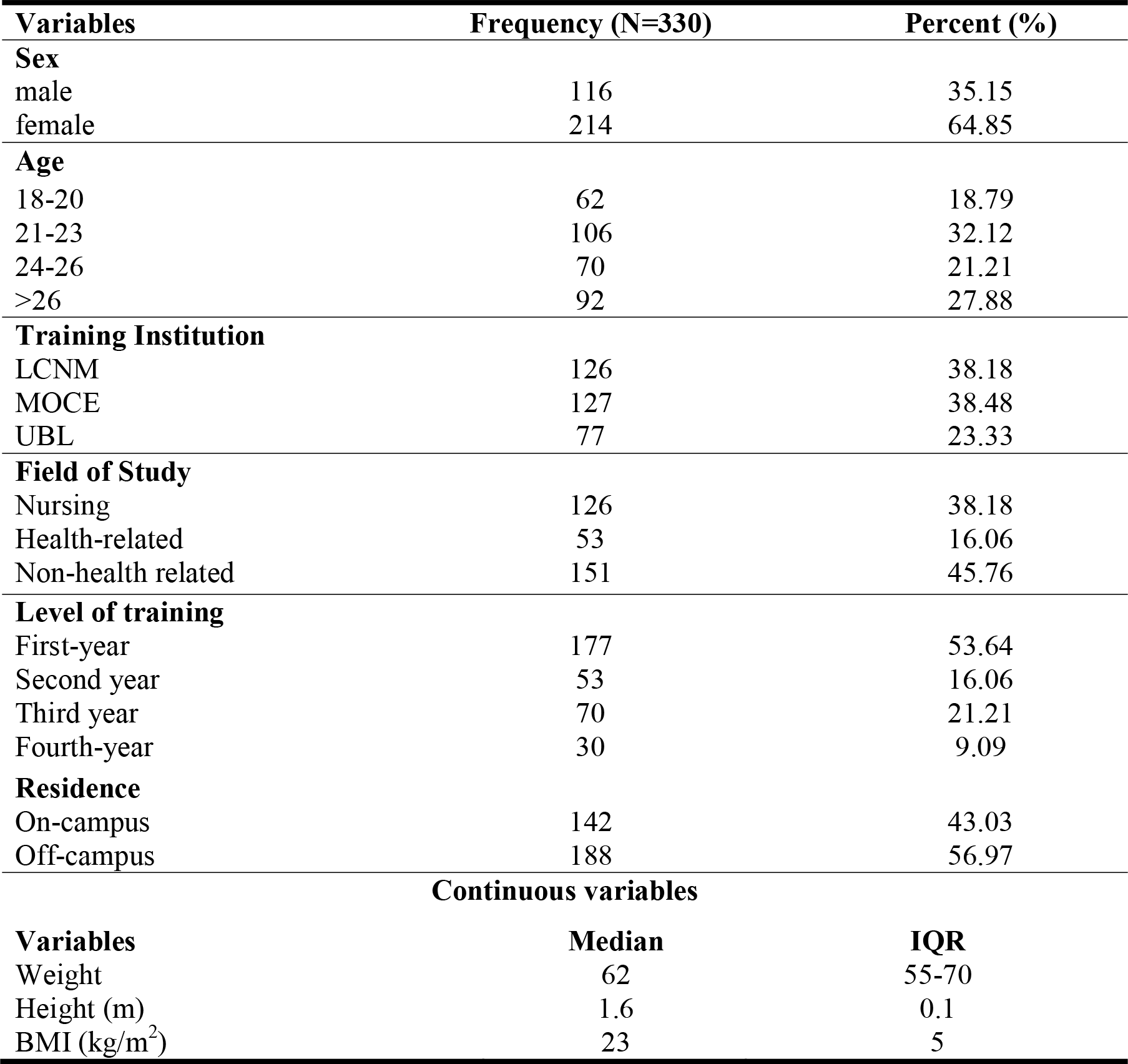
Socio-demographic profile and continuous variables of respondents.

### Overweight and Obesity Prevalence

The overall prevalence of overweight and obesity among respondents was 30.6% (23.0% overweight and 7.6% obese). Female respondents 70 (69.3%) were significantly more overweight and obese than male respondents 31 (30.7%). Concerning the field of study, a higher prevalence of overweight and obesity was found among non-health-related students, 47 (46.5%) compared to nursing students, 29 (28.7%) and health-related students, 25 (24.8%).

### Field of study

Table 2 reveals that students’ field of study was significantly associated with overweight and obesity among respondents (P=0.003). In addition, the results in Table 5 show that students from a non-health-related field recorded the highest proportion of overweight and obesity 47/101 (46.53%) compared to students from a nursing field 29/101 (28.71%), and a health-related field 25/101 (24.75%) (P=0.003).

**Table 2:** Association between the field of study and BMI.

| BMI | nursing | Health-related | Non-health related | P-value* |
| --- | --- | --- | --- | --- |
| <18.5kg/m<sup>2</sup> | 14 | 1 | 6 | 0.003 |
| 18.5-24.9kg/m<sup>2</sup> | 83 | 27 | 98 |  |
| 25-29.9kg/m<sup>2</sup> | 25 | 16 | 35 |  |
| ≥30 kg/m<sup>2</sup> | 4 | 9 | 12 |  |
| Total | 126 | 53 | 151 | 330 |

\*Chi-squared Test

### Food Consumption Patterns

Table 3 shows the feeding patterns and food sources of overweight/obese respondents in the study. A high percentage of 33 (40.74%) of the respondents reported consuming snacks once every day. In comparison, 39 (38.61%) of the respondents reported consuming fruits and vegetables once every day and (36.36%) of the respondents consumed fast food once every day (Table 3). A total of 83 (35.62%) of respondents reported consuming junk food once per week, while 234 (71.12%) reported consuming nshima twice per day.

**Table 3:** Food consumption pattern of respondents (n=101).

| Variables | Frequency | Percent (%) |
| --- | --- | --- |
| **Fruits and vegetables** |  |  |
| Once every day | 39 | 38.61 |
| 2-3 times a day | 33 | 32.67 |
| Twice in a week | 19 | 18.81 |
| Once in a week | 10 | 9.90 |
| **Total** | 101 | 100 |
| **Snacks** |  |  |
| Once every day | 33 | 40.74 |
| 2-3 times a day | 11 | 13.58 |
| Twice in a week | 17 | 20.99 |
| Once in a week | 20 | 24.69 |
| **Total** | 81 | 100 |
| **Fast food** |  |  |
| Once every day | 28 | 36.36 |
| 2-3 times a day | 9 | 11.69 |
| Twice in a week | 18 | 23.38 |
| Once in a week | 22 | 28.57 |
| **Total** | 77 | 100 |
| **Junk food** |  |  |
| Once every day | 59 | 25.32 |
| 2-3 times a day | 38 | 16.31 |
| Twice in a week | 53 | 22.75 |
| Once in a week | 83 | 35.62 |
| **Total** | 233 | 100 |
| **Nshima** |  |  |
| Once every day | 65 | 19.76 |
| Twice per day | 234 | 71.12 |
| Three times per day | 30 | 9.12 |
| **Total** | 329 | 100 |

### Alcohol intake

Cross tabulation and chi-square test revealed a significant association between alcohol consumption and overweight or obesity (P=0.003) (Table 4). More students did not take alcohol compared to those who took alcohol, i.e. 245 and 85, respectively.

**Table 4:**
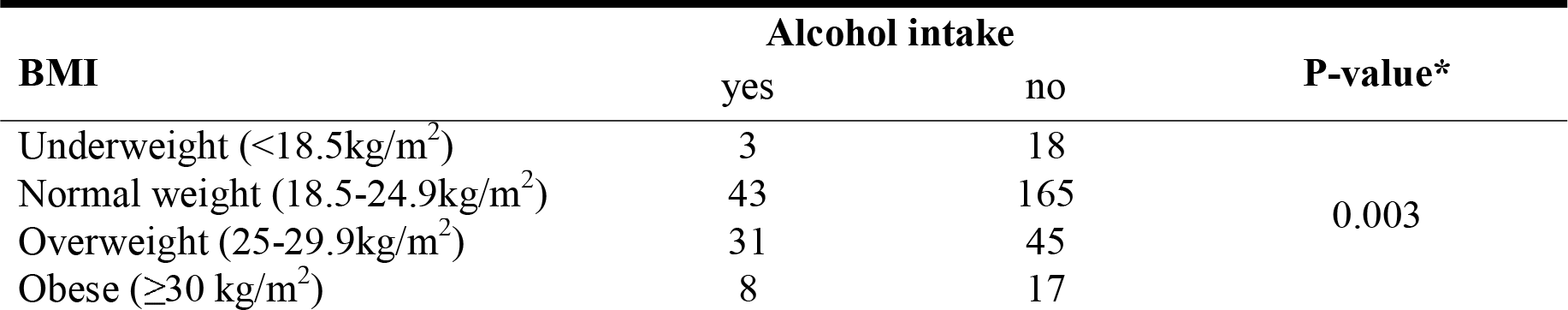
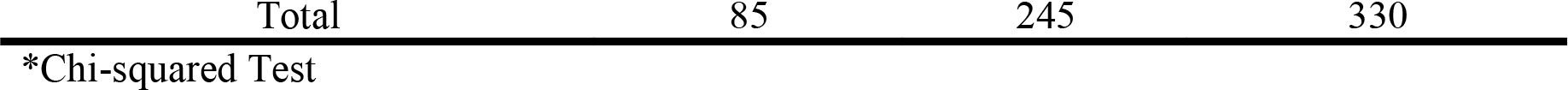
Association between alcohol intake and BMI.

| BMI | Alcohol intake |  | P-value* |
| --- | --- | --- | --- |
|  | yes | no |  |
| Underweight ($<18.5\text{kg/m}^2$) | 3 | 18 | 0.003 |
| Normal weight ($18.5\text{-}24.9\text{kg/m}^2$) | 43 | 165 | |
| Overweight ($25\text{-}29.9\text{kg/m}^2$) | 31 | 45 | |
| Obese ($\geq 30\text{ kg/m}^2$) | 8 | 17 | |

\*Chi-squared Test

### Association between independent Variables and dependent variables

The independent variables significantly associated with overweight and obesity among the participants were age group older than 26 years among the students, which had reduced odds 0.37 (95%CI 0.18 – 0.79) compared to students aged between 18 to 20 years. Sex was also statistically significantly associated with being overweight and obese; Female students were less likely to be overweight and obese with an adjusted odds ratio of 0.59 (0.18 – 0.79) compared to their male counterparts after adjusting for age and behavioural factors such as exercises, food and alcohol intake patterns. The other variable associated with overweight and obesity was alcohol intake, which increased the odds by 1.51 aOR 2.51 (1.44 – 4.38). Table 5 shows the associations of selected factors and the outcome variable:

**Table 5:**
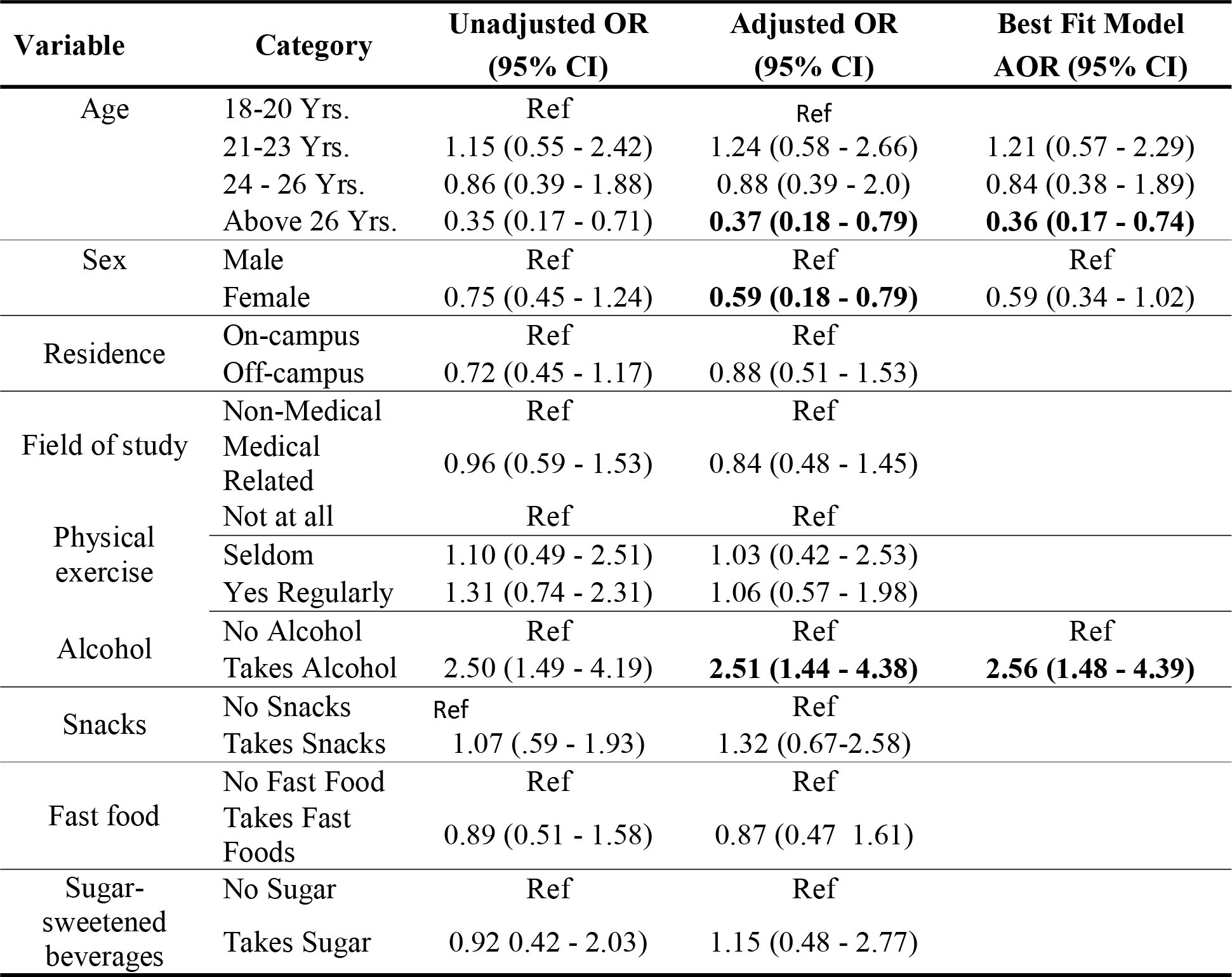
Association between independent and dependent variables.

### Model Selection Criteria

Based on the Akaike and Bayesian information, model fitness was assessed for the lowest and highest levels of the coefficient of determination (Pseudo R^2^). The model with age, sex and alcohol had the lower AIC and BIC but reduced coefficient of determination compared to the full model as shown in table 6. We, therefore, adopted the full model as our best-fit model as it was better able to explain the variation in the outcome despite having higher values of AIC and BIC.

**Table 6:** Model Selection Criteria.

| Model | Pseudo R-Squared | AIC | BIC | P-Value |
| --- | --- | --- | --- | --- |
| Full model (Age, sex, Residence, field of study, physical exercise, alcohol, fruits and vegetables, fast food, junk food, and sugar-sweetened beverages). | 0.082 | 401.30 | 454.49 | 0.002 |
| Reduced Model(Age, sex, and alcohol). | 0.078 | 386.92 | 409.71 | <0.001 |

\*The Akaike Information Criteria (AIC) and Bayesian (BIC) Information Criteria

## DISCUSSION

The current study was conducted to determine the prevalence of overweight and obesity among college students and the associated risk factors. The study has shown that over a fifth of the students were overweight, and at least one-third were overweight or obese. While this is not representative of the general population, it is concerning that a third of young adults in colleges are either overweight or obese. A study that used nationwide data on women in 2014 found a prevalence of 26.4%, which is comparable to this study’s findings of 30.6%, although the former only included women (Bwalya *et al*., 2017). Furthermore, these results correlate with findings from the study carried out on food consumption patterns, physical activity and overweight and obesity among undergraduates of a private university in Nigeria, which showed that 31% of respondents were overweight and 9.3% were obese (Kayode *et al*., 2020).

This study revealed that female sex reduced the odds of being overweight or obese (AOR=0.59; 95% CI=O.18 - 0.79). This is consistent with studies done by Jiang *et al*. (2018) on overweight and obesity among Chinese college students and a study done in Brazil among nursing students by Urbanetto *et al*. (2019), which indicated that males were more susceptible to being overweight and obese than females.

Our study found that older age groups above 26 years had reduced odds of obesity compared to the younger age group of 18 to 20 years old. This may be due to the fact that older age groups are more assertive and able to take care of their habits, such as alcohol and junk food. A study done among health sciences college students in Saudi Arabia found that age was not associated with obesity (Makkawy *et al*., 2021). Our study found that regular alcohol intake was associated with overweight and obesity. This is consistent with findings from a systematic review conducted by Traversy *et al*. (2015), which showed a positive association between alcohol consumption and measures of abdominal adiposity (also known as “beer belly”).

This study did not find significant associations between overweight and obesity and other variables such as physical exercises, taking of snacks, fast foods and sugar-sweetened beverages. Our study may not have been powered to detect differences for all the variables; regular exercise has been shown to reduce weight (Tateyama, 2019). Other studies have also shown that taking snacks, fast foods and sugar-sweetened beverages is associated with overweight and obesity (Moise, 2019).

### Study limitations

Anthropometric measurements only included height, weight and body mass index (BMI). Since BMI is not the only accurate measure for adiposity, waist circumference and waist-to-hip ratios should be incorporated in future research. Secondly, this is a cross-sectional study, so it does not adduce causality but associations.

### Conclusion

This study has found a high prevalence of overweight and obesity among young adults attending college education. Sex, age, and alcohol intake were significantly associated with overweight and obesity among students.

## Data Availability

All data is available upon request to the corresponding author.

